# An exploration of the genetics of the mutant *Huntingtin* (*mHtt*) gene in a cohort of patients with chorea from different tribes in 6 sub-Saharan African countries

**DOI:** 10.1101/2022.07.13.22272435

**Authors:** Mendi J Muthinja, Carlos Othon Guelngar, Maouly Fall, Fatumah Jama, Huda Aldeen Shuja, Jamila Nambafu, Abdoulaye Bocoum, Daniel Gams Massi, Oluwadamilola O. Ojo, Njideka U. Okubadejo, Funmilola Tolulope Taiwo, Alassane Mamadou Diop, Coudjou J.D.G. De Chacus, Fodé Abass Cissé, Amara Cissé, Juzar Hooker, Dilraj Sokhi, Henry Houlden, Mie Rizig

**Author notes:** Correspondence to Dr. Mie Rizig, Department of Neuromuscular Diseases, UCL Queen Square Institute of Neurology, London WC1N 3BG, UK, Dr. Mendi J. Muthinja, Huntington Disease Africa, P.O. Box. 46092-00100, Nairobi, Kenya.

## Abstract

**Background:** Africans are underrepresented in Huntington’s disease research. A European ancestor was postulated to have introduced the mutant *Huntingtin* (*mHtt*) gene to the continent, however recent work has shown the existence of a unique *Huntingtin* haplotype in South-Africa specific to indigenous Africans.

**Objective:** We aimed to investigate the CAG repeats expansion in the *Huntingtin* (*Htt*) gene in a geographically diverse cohort of patients with chorea and unaffected controls from sub-Saharan Africa.

**Methods:** We evaluated 104 participants: 45 patients with chorea, 24 asymptomatic first-degree relatives of subjects with chorea and 35 healthy controls for the presence of the *mHtt*. Participants were recruited from 6 African countries. Additional data were collected from patients positive for the *mHtt* including demographics, presence of psychiatric and cognitive symptoms, family history, spoken languages and tribal origin. Additionally, their pedigrees were examined to estimate the number of people at risk of developing HD and to trace back the earliest account of the disease in each region.

**Results:** HD cases were identified in all countries. 53.3% of patients with chorea were carriers for the *mHTT*; median tract size 45 CAG repeats. Of the asymptomatic relatives 41.6% were carriers for the *mHTT;* median tract size 42.5 CAG. No homozygous carries were identified. Median CAG tract size in controls was 17 CAG repeats. Men and women were equally affected by HD. All patients with HD—bar three who were juvenile onset of <21 years—were defined as adult onset (median age of onset 40 years). HD transmission followed an autosomal dominant pattern in 80% (16/20) of HD families. In familial cases, maternal transmission was higher (56%) than paternal transmission (44%). The number of asymptomatic individuals at risk of developing HD was estimated at 10 times more than the symptomatic patients. HD could be traced back to the early 1900s in most African sites. HD cases spread over 8 tribes belonging to two distinct linguistic lineages separated from each other approximately 37 kya ago: Nilo-Sahara and Niger-Congo.

**Conclusion:** This is the first study examining HD in multiple sites in sub-Saharan Africa. We demonstrated that HD is found in multiple tribes residing in 6 sub-Saharan African countries including the first genetically confirmed HD cases from Guinea and Kenya. The prevalence of HD in the African continent, its associated socio-economic impact, and genetic origins need further exploration and reappraisal.

## Introduction

Huntington’s disease (HD) is an autosomal dominant neurodegenerative condition causing progressive loss of cognition, movement disorder, and psychiatric symptoms^1^. HD results from an expansion of a CAG nucleotides repeat within exon 1 of the *Huntingtin (HTT)* gene^2,3,4^.

Historically, HD was first described by the name “setesdalryyja” in 1860 by the Norwegian neurologist Johan Lund^5^. Twelve years later, George S Huntington published his famous assay “On Chorea” in North America, describing the clinical manifestation of the disease in English^1^. Since then HD cases have been reported in most countries across the globe^1,6-9^. In Africa, the earliest documented account of HD was in 1936 in a Mugikuyu psychiatric patient from Kenya^10^. In 1981, Scrimgeour, reported a family with several affected individuals with possible affected ancestors that could be traced back to at least two people born in the 1870s, in Tanzania^11^. Elsewhere in Africa, clinical accounts of HD have been reported from Nigeria, Senegal, South Africa, Sudan, Tanzania, Togo, Uganda and Zimbabwe^11-17^. Genetic testing was used in the reporting of cases positive for *Huntingtin* mutation (*mHTT)* in Burkina Faso, South Africa, Egypt, Morocco, Mali and the Gambia^18-23^. However, the knowledge about the exact origin and the distribution of *mHTT* in the continent is sparse. A small number of studies traced the origin of the *mHTT* to ethnic groups from South Africa, Tanzania, and Zimbabwe, and suggested that the mutation was either introduced by European migrations or originated from native pathogenic variants in the *HTT* gene as demonstrated by haplotype analyses^24-28^.

Novel therapeutic approaches targeting the *mHTT* are currently in clinical trial^29,30^. Addressing gaps in knowledge about HD among sub-Saharan Africans is needed if people of this ethnic origin are to benefit from developing novel therapies. To achieve this objective, we formed a consortium—the African Research Consortium on Huntington’s disease (ARCH)—comprising of predominantly early career African academics and movement disorders specialists whose focus is studying HD in the continent. We aim to characterize genetically and phenotypically a geographically diverse cohort of cases of HD from sub-Saharan Africa. Here we report on ARCH’s first cohort study from 6 African countries.

## Materials and Methods

Patients with chorea and their first-degree relatives were recruited from movement disorder clinics in 6 African countries: Cameroon, Guinea, Kenya, Mali, Nigeria, and Senegal, between January 2020 and October 2021 (Figure 1). Cases were identified from medical records at the participating centres. Healthy individuals without a neurological illness or a family history of a neurological illness from the same populations were included as controls. Participant selection and assessments were performed by movement disorders specialists authoring this manuscript.

**Figure 1:**
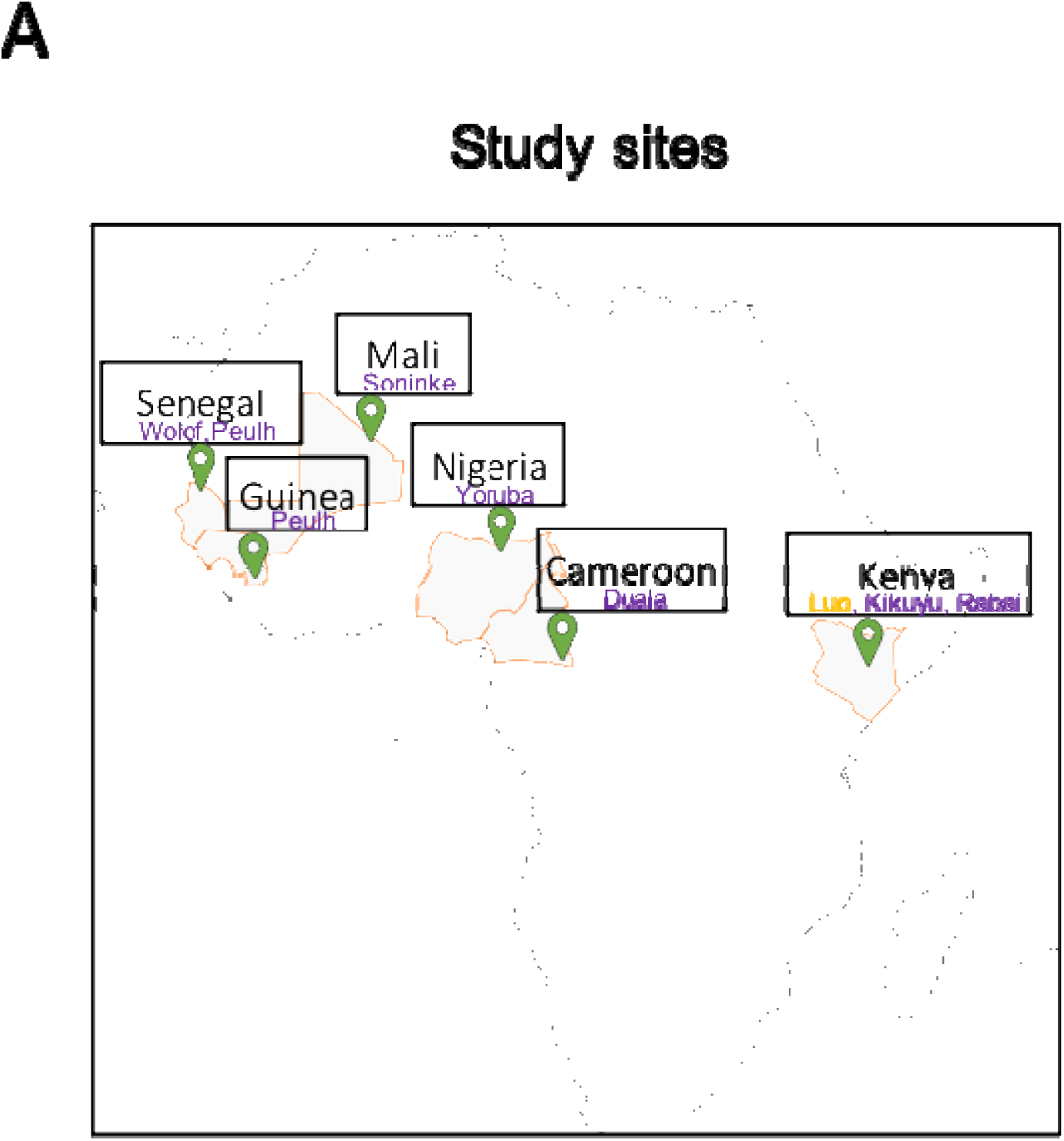
Study sites and tribal origins of HD families. Movement disorders centres in 6 African countries contributed to this cohort. In total 20 HD families from 8 tribes were identified. These tribes are belonging to two ethnolinguistic origins - a Niger-Congo (purple) and Nilo-Saharan (yellow). The 8 tribes were listed in the map above according to their locations.

Ethical approval was obtained from the institutional research ethics boards of each of the six African countries and the ethics committee of the University College London and the National Hospital for Neurology and Neurosurgery, London, UK. All participants gave written informed consent at each study site according to the Declaration of Helsinki and The Common Rule. Consents covered participation in research, publications and data sharing with bona fide researchers. For individuals deemed to lack capacity to consent, study sites applied country-specific guidelines for signing consent forms. Minors were included only with consent from both parents.

DNA was isolated from either EDTA-blood or buccal swabs or saliva. Fragments size analysis using a PCR method followed by capillary electrophoresis was applied to estimate the CAG tract size in exon 1 of the *HTT* gene, as previously described^31^. All samples except those from Kenya were genotyped at UCL Queen Square Institute of Neurology, London, UK. The Kenyan samples were genotyped either commercially or in-house at the Aga Khan University Hospital in Nairobi.

Additional data from individuals with the *mHTT* (CAG tract >35) were collected; this included gender, any behavioural, psychiatric and cognitive symptoms, age at experiencing the first neurological or behavioural and psychiatric manifestations, family history, spoken language, and tribal origin.

Genealogical analysis was performed by examining at least three generations for each individual’s family to acquire sufficient information on consanguinity, number of individuals in each generation, number of individuals with possible HD symptoms, and the age when symptoms were experienced, and age at death (if deceased). Whenever possible, family trees were extended back to the fourth and fifth generations to trace back the oldest known individuals with HD symptoms. Additionally, pedigrees were examined to estimate the transmission pattern (i.e., maternal or paternal) of the *mHTT* and the total number of living persons at risk of developing HD. Data analysis was performed using Microsoft Excel or GraphPad Prism Version 8.0.

## Results

CAG tract sizes were determined from 104 participants: 45 patients with chorea, 24 asymptomatic first-degree relatives and 35 unrelated healthy controls. 191 out of possible 208 alleles were successfully genotyped; 17 alleles dropped out during genotyping due to PCR failure (Figure 2A-C).

**Figure 2:**
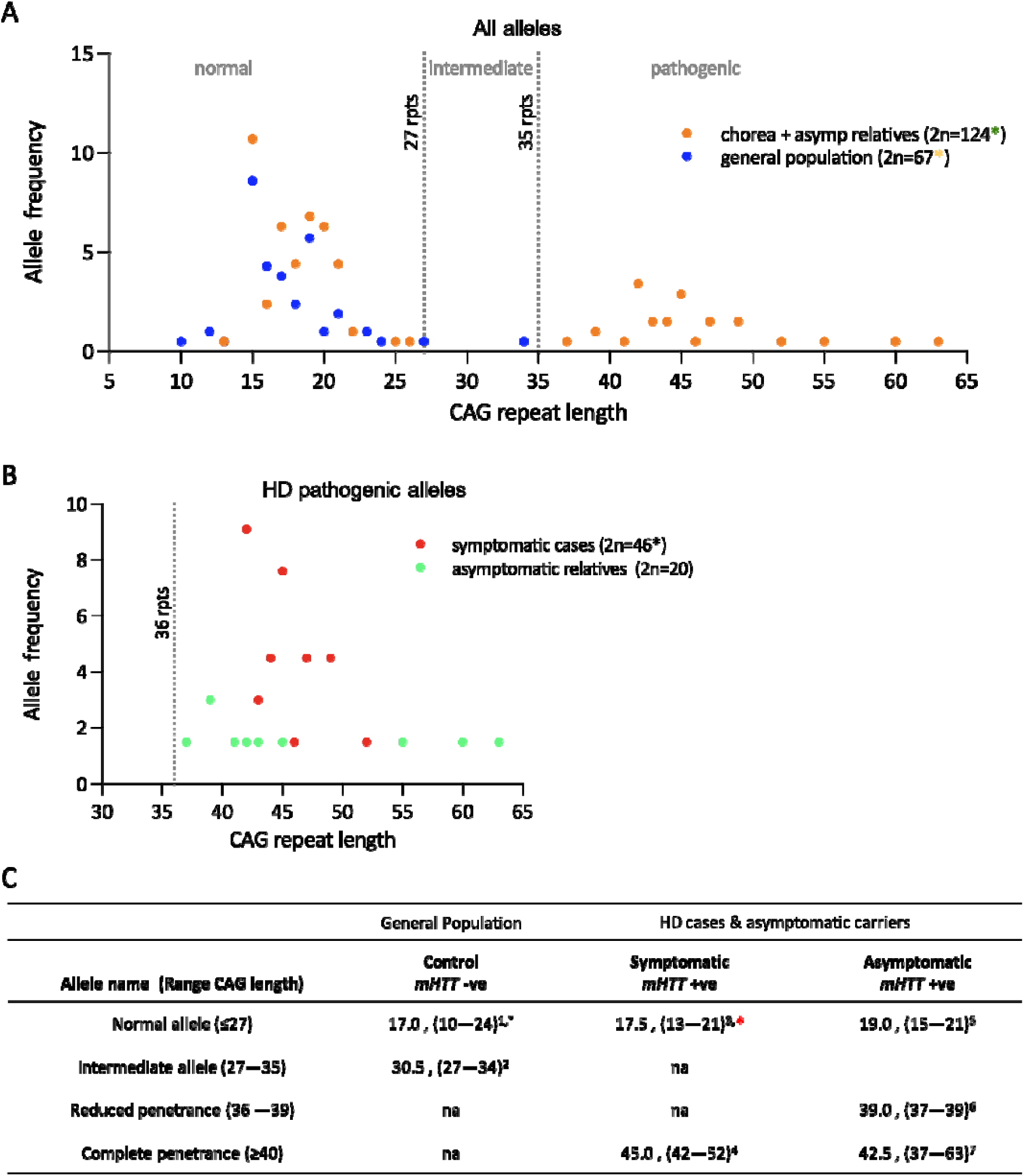
Allele Frequency distribution. (A) Allele frequencies of CAG repeats in all 104 participants (i.e., 45 patients, 24 asymptomatic relatives and 35 controls). CAG tracts are categorized as normal (wild type), intermediate or pathogenic. Allele dropouts due to PCR failure are denoted as asterisks: black or red asterisk =2 alleles, amber asterisk= 3 alleles and green asterisk= 14 alleles. (B) Allele frequencies of expanded-pathogenic CAG repeats (>35), in patients and asymptomatic relatives who are positive for the HD gene. (C) Alleles’ classification for controls, patients and asymptomatic carriers. Superscripts represent the number of alleles where: ^1^(2n=64), ^*^(3 ADO); ^2^(n=2); ^3^(n=22), ^*^(2 ADO); ^4^(n=24); ^5^(n=10); ^6^(n=3); and ^7^(n=7).

The size of the most prevalent wild type allele in all individuals was 15 repeats (cumulatively at 8.6%), and the size of the most prevalent *mHTT* in patients and asymptomatic relatives was 42 repeats (cumulatively at 3.4%). In total 34 individuals (24 patients and 10 asymptomatic relatives) carried one allele with CAG repeats within the pathogenic range for HD (>35 repeats) (Figure 2A-C). No homozygous carriers were identified.

53.3% (24/45) of patients were heterozygous carriers for the *mHTT* allele; with median repeat sizes of 45 (mode 42, range 42–52). 41.6% (10/24) of the asymptomatic relatives were heterozygous carriers for the *mHTT* allele; with a median repeat size of 42.5 (mode 39, range 37—63). In controls the median CAG tract size was 17 (mode 15, range 10—34). Notably, all 24 patients who were carriers of the *mHTT*, had expanded CAG repeats within the complete penetrance category of >39 repeats. Intermediate CAG repeats with lengths measuring 27 and 34 were detected only in two controls—one from Guinea and one from Senegal respectively. The tract sizes and the frequency distributions of the wild type *HTT* (< 27 CAG repeats), intermediate *HTT* (27–35 CAG repeats) and the *mHTT* allele (>35 CAG repeats) in all individuals, are summarised in Figure 2 (A-C).

The 24 patients with the *mHTT* belonged to 20 distinct families that were identified from all the six recruitment sites: Cameroon 1 patient (1 family), Guinea 5 patients (5 families), Kenya 5patients (5 families), Mali 1patient (1 family), Nigeria 4 patients (2 families), and Senegal 8 patients (6 families). Men and women were equally affected by HD (12 out of 24 each). 79% (19/24) HD patients had cognitive disturbance and 75% (18/24) had behavioural and psychiatric symptoms. 87.5% (21/24) developed adult-onset HD (age in years: median 40, mode 45, range 23—62). Juvenile onset HD affected 12.5% (3/24) of patients; of these cases, 2 were from Senegal (aged 18 and 20 years) with 46 and 45 CAG repeats respectively. A single case of a 20-year-old juvenile HD with 49 CAG repeats was identified from Cameroon.

A multigenerational history of HD symptoms was identified in 80% (16/20) of the families. 4 out of the 20 families were without a multigenerational HD history; one proband from Kenya was a confirmed sporadic case of HD and in 3 families, from Cameroon, Mali and Senegal, there was insufficient genealogical data to infer a clear family history of HD. Of the families with a clear multigenerational history of HD, 56% (9/16) had a maternal transmission while 44% (7/16) had a paternal transmission pattern. 10% (2/20) of families had consanguineous marriages: 1 from Guinea and 1 from Senegal.

An additional 15—21 males and 6 females—living individuals were identified as possibly having symptomatic HD in 2 to 4 generations in the 20 families. A further 200 were estimated to be asymptomatic and at-risk for HD (110 males, 87 females, and 3 unknown). Note that, at risk individuals were asymptomatic, and of undetermined HD genotype, but had shared ancestry with the proband over several generations. Of the 200 persons at risk, 135 were first degree relatives i.e., parents, offspring or siblings of the proband. The largest number of people at risk were from large families in Guinea, Kenya, and Senegal.

A possible history of HD—based on tracing the oldest HD ancestors with symptoms in the 20 families—was traced back to the 1900s at most sites. In at least one large kindred from Kenya we have been able to trace accounts of HD to the early 1900; with possible relatives stretching back to the mid-1800s (Supplementary Figure A).

HD patients were linked to 8 genetically unique tribal groups: Fulani (Peulh), Wolof, Soninke, and Yoruba from West Africa; the Duala from Central Africa and the Rabai, Kikuyu and Luo from East Africa (Figure 1). The 8 tribes came from 2 distinct ethnolinguistic origin: Niger-Congo lineages included Fulani (Peulh), Wolof, Soninke, Yoruba, Duala, Rabai and Kikuyu while the Luo people were from a Nilo-Saharan lineage. 95% (19/20 families) were identified as Niger-Congo and only 5% (1/20 families) were identified as Nilo-Saharan (Figure 1). All date produced in this study including family trees and detailed regional observations of HD patients and their families per country could be provided to bona fide researchers upon reasonable written request to ARCH study group.

## Discussion

In Kenya, in 2020, the Swahili word “Uku” was designated by Huntington disease Africa (https://hd-africa.org/) to mean HD. In the majority of the local communities in Africa HD is not recognized and it has no name. Internationally, little is known about HD in Africans^32,33^. Current knowledge comes from a few random case reports and several South African-based genetic studies mainly in people from a mixed ancestry or Bantu-speaking tribes^20-23,27,33,34^. Here we report on a cohort of African patients and first-degree relatives with a confirmed genetic diagnosis of HD from 8 different tribal origins spanning 6 sub-Saharan countries—including the first genetically confirmed HD cases from Guinea and Kenya.

53% of patients with chorea in this cohort tested positive for the *mHTT*; this percentage is higher than previously reported in black South Africans (36%), but lower than the reported figures from other populations^33^. It is plausible that a significant proportion of the 47% of patients in our study who are negative of the *mHTT* to be harbouring HD phenocopies such as the Huntington’s disease-like syndrome (HDL) 2; which was reported solely in patients with chorea from black populations; further characterisation of these HD negative cases is required^33^.

The *mHTT* tract sizes in our patients and their asymptomatic relatives were similar to the previously reported sizes in both black South Africans^27,33^ and in HD cases in other ethnic groups^25^, however, several observations from our study — and in line with previous studies — suggest that the *HTT* locus in indigenous sub-Saharan African populations is possibly more stable than in Europeans^35^. Firstly, the commonest CAG repeat size in our cohort was 15, similar to previous reports in black south Africans which is less than the average tract sizes in Europeans (18) and Asians (17)^36,37^. Secondly, all HD cases in our study had CAG tracts within the full penetrance range (>39 repeats), comparable to reports from black south Africans where 98-99% of black patients with HD had repeats sizes within full penetrance range^27,33^. Thirdly, in our cohort the CAG repeats within the reduced penetrance range (36-39 repeats) were only detected in asymptomatic relatives and none of the controls had alleles within the mutable range.

With few regional variations, most HD patients in our study developed symptoms in their thirties and forties; the eldest patient being 62 years old. Psychiatric and cognitive symptoms were identified in over 70% of HD patients: slightly more than reported in other studies^38^. Juvenile cases constituted 12% (3/24) of HD patients, however due to the small size of this cohort it is difficult to draw comparisons between this percentage and the 4-5% rate reported in European and North American cohorts^39,40^. Notably however, the clinical presentation and the tract sizes of our juvenile cases did not differ from the adults with HD.

As expected with an autosomal dominant pattern of inheritance, 80% of our families had a multigenerational history of HD symptoms and 40% of the first-degree asymptomatic relatives included in this study were found to be carriers of the *mHTT*. Consanguinity was detected in 10% (2/20) of HD families; highlighting the need to explore the role of customary and traditional practices in hereditary rare diseases in these indigenous communities. In familial cases, maternal transmission was higher than paternal transmission (55% and 46% respectively). No gender differences were observed. Nonetheless, inspection of the family trees of affected individuals showed a higher proportion of males are at risk of carrying the *mHTT*, compared to females. The number of individuals identified at risk of developing HD was estimated to be around 10 times the number of people tested positive in our study; this raises the possibility of a higher prevalence of HD across sub-Saharan African than currently believed^14,32^.

One of the important findings from this cohort is that in one of the Kenyan families, we were able to trace back an ancestor with HD symptoms to the early 1900 (Supplementary figure A). This account suggests that the preceding generation, of the reported family could be from a similar time point as the previously reported earliest case in the literature by Scrimgeour in 1870s^11^ and is possibly one of the two earliest documented accounts in Black Africans. Another important finding is that the patients with *mHTT* allele were widely spread across 8 tribes in sub-Saharan Africa— belonging to two genetically distinct ethnolinguistic lineages (the Niger-Congo and Nilo-Sahara) which diverged approximately ∼□16 kya ago^41^—suggesting that perhaps the present-day ancestors carrying *mHtt* from these lineages had multiple ancestral origins for the CAG expansion mutation. The multiple ancestral origins possibly coincide with migratory routes of modern-day Sub-Saharan Africans challenging the theory that the mutation was first brought to the continent by Europeans. Indeed, investigating the *HTT* haplotypes in these distinct tribes is essential to trace the origin of *mHTT* in sub-Saharan Africa.

In conclusion, we have identified sub-Saharan African families with *mHTT* from different tribes across the region. This cohort is the largest group of patients with genetically confirmed HD from different countries in the continent. Our findings call for a revision of the possibly underestimated HD prevalence data in Africa. Further investigation of the genetic variations, haplotypes, and modifiers of HD progression for this cohort are also warranted. We acknowledge the limitations of this study— including the small sample size, the lack of detailed clinical assessment using standardised rating scales for HD patients.

Planned future work from our group includes conducting detailed phenotypic assessments of this cohort using clinical scales and imaging. To better understand the HD locus in Africans we are planning to assess haplotypes and within tract changes in patients with HD and to investigate HD photocopies in HD negative cases. In collaboration with Huntington disease Africa (https://hd-africa.org/) — the main HD-patient advocacy group in Africa — our ARCH group aims to build a registry for HD featuring patients and asymptomatic relatives from the 6 African participating countries. We hope this registry will serve as a foundation for epidemiology, clinical, biomarkers and genetic studies as well as a base to collaborate with international efforts such as Enrol-HD study.

## Data Availability

All data produced in the present study are available upon reasonable request to the authors

## Supplementary materials

**Supplementary material A:**
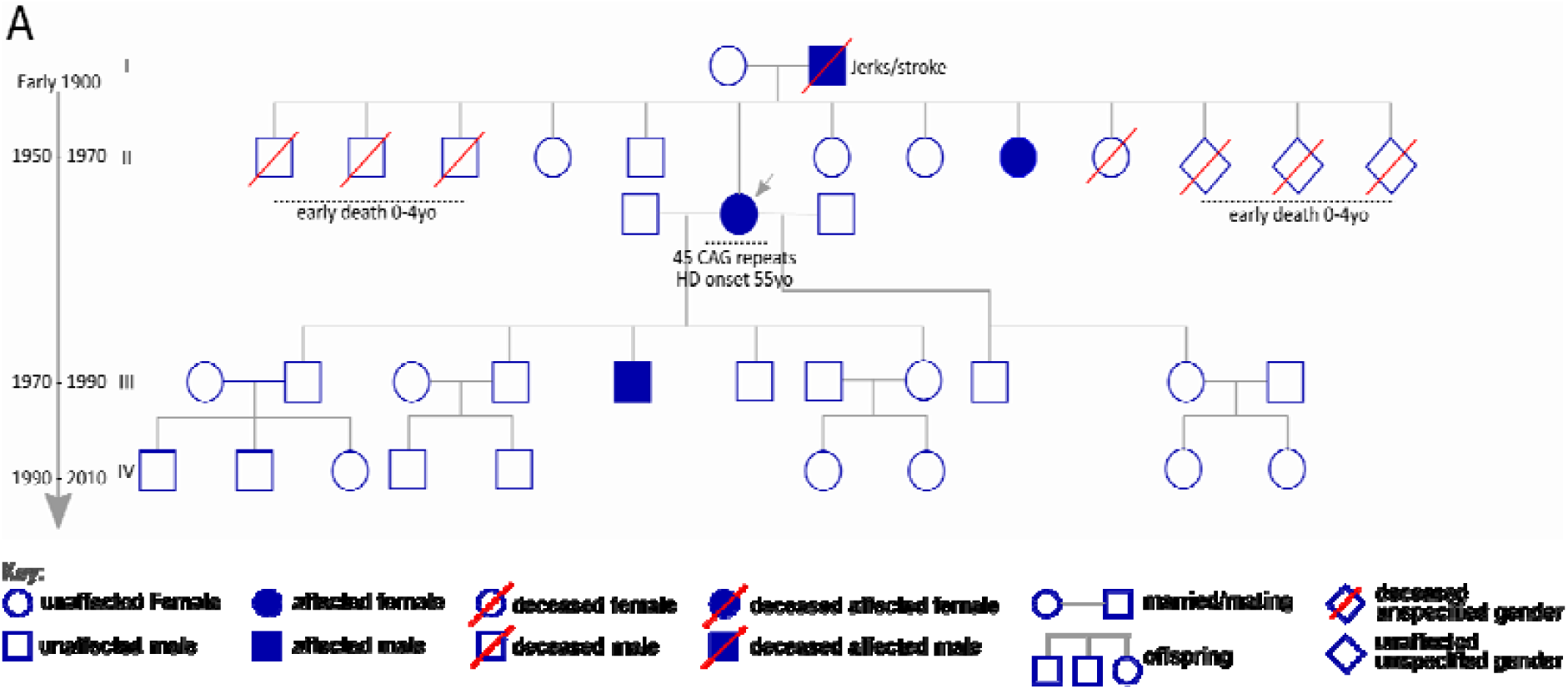
A family tree of a multigenerational Kenyan family with >120 years HD history: A pedigree of a multigenerational Kenyan HD family that was clinically examined and also genetically tested for presence of the HD pathogenic allele. The offspring is arranged in ascending order, from the youngest to eldest child. Grey arrow indicates proband. Arrow on the side indicate generational estimated date of births.

